# eSexualHealth: Preferences to Use Technology to Promote Sexual Health Among Men Who Have Sex with Men and Trans and Gender Diverse People

**DOI:** 10.1101/2022.09.26.22280388

**Authors:** Esha Abraham, Eric PF Chow, Christopher K Fairley, David Lee, Fabian YS Kong, Limin Mao, Jane L Goller, Nicholas Medland, Benjamin R Bavinton, Budiadi Sudarto, Stefan Joksic, Jessica Wong, Tiffany R Phillips, Jason J Ong

## Abstract

**Objectives:** Gay, bisexual and other men who have sex with men (GBMSM) and trans and gender diverse (TGD) people are disproportionately affected by poorer sexual health outcomes compared to heterosexual populations. We aimed to explore the preferences of GBMSM and TGD for using eHealth for sexual health (eSexualHealth).

**Methods:** We distributed an anonymous online survey among the lesbian, gay, bisexual, transgender, intersex, queer and other people of diverse sexuality or gender (LGBTIQA+) community in Australia. The survey collected data on sociodemographic characteristics and sexual behaviours, their preferences for app/website functions and preferred HIV and sexually transmitted infection (STI) testing reminders. We used descriptive statistics to summarise the characteristics of the study population. Free-text responses were thematically analysed.

**Results:** Of 466 participants included, most identified as cisgender males (92.7%). The median age was 48 (interquartile range [IQR]: 37-56). For accessing sexual health-related information, 160 (34.6%) would use either a website or an app, 165 (32.7%) would prefer a website, 119 (25.8%) would prefer an app, and 33 (7.1%) would not use either platform. There was no significant difference between GBMSM and TGD people. Participants were most interested in information about STI clinics, HIV/STI hotspots, and sexual health education. Participants stressed the need for privacy and anonymity when using eHealth. Regarding reminders to test for HIV/STIs, receiving regular SMS was most popular (112/293, 38.2%), followed by regular emails (55/293 18.8%) and a reminder function on their phone (48/293, 16.4%).

**Conclusion:** Our study suggests a promising future for eHealth among GBMSM and TGD people. Sexual health is still a stigmatised area, and eHealth may circumvent barriers this population faces.

**What is already known about the topic:**

- Gay, bisexual, and other men who have sex with men (GBMSM) and trans- and gender diverse people (TGD) have poorer sexual health outcomes compared to heterosexuals
- eHealth (or digital interventions) to improve sexual health is a growing area

**What this study adds:**

- Our survey among Australian GBMSM and TGD individuals document their preferences for using eHealth to optimize their sexual health

**How this study might affect research, practice or policy:**

- Being aware of preferences for eHealth can inform resource allocation and future development of features wanted by target populations

## Introduction

While HIV notification rates have decreased in Australia in recent years, gay and bisexual men who have sex with men (GBMSM) continue to be disproportionally affected compared to their heterosexual counterparts.^1^ Furthermore, STI incidence rates are higher among GBMSM living with HIV than among HIV-negative GBMSM.^2^ A 2018 survey of trans and gender diverse (TGD) people found that more than half believed they received poor sexual health education during their school years. Additionally, 51.2% reported receiving insensitive sexual health care. Most participants (65%) also reported inconsistent condom use with casual sexual partners. Combined with their poor experiences in sexual health care, it increases their risk for HIV/STIs.^3^ As a result, greater effort needs to be put into educating and facilitating HIV/STI testing among GBMSM and TGD people.

GBMSM and TGD people can face several barriers when accessing healthcare, such as: experiencing stigma due to their sexuality, a lack of knowledge and culturally appropriate training amongst healthcare providers; and personal concerns around disclosing their sexual identity.^4-6^ Specialised sexual healthcare is scarce in Australia, and patients can face long waiting times or high costs. Digital health interventions, also known as eHealth, can improve healthcare access among LGBTIQA+ people.^7^ These can be delivered through mobile devices, laptops, websites, or smartphone apps, and can provide private, personalised content that is easily accessible.^8^ However, to be successful, input from the target audience is essential in creating an intervention that addresses their needs and, in turn, results in increased uptake.^9 10^ Previous studies have determined that there is a promising future for an eHealth app among GBMSM.^11^ However, few studies have investigated the preferences and types of features for different platforms.

This study sought to understand the features of a website or smartphone app that GBMSM and TGD people prefer to access information related to sexual health. We also aimed to measure whether GBMSM and TGD people currently use eHealth for reminding themselves to test for HIV/STIs.

## Method

### Study Population

This was an anonymous online survey distributed among the LGBTIQA+ community in Australia. We included all respondents aged 16 years and above who identified as LGBTQIA+. Completion of the survey was taken as implied consent. The online survey link was disseminated through the authors’ professional networks, social media, and clients at Melbourne Sexual Health Centre (MSHC), a public sexual health clinic in Australia. This included a short messaging service (SMS) or email from MSHC (if they had previously consented to receive them), a dating app (Grindr), and LGBTIQA+ community groups. These community groups included Equinox, Your Community Health, Switchboard, Minus18, QLife, and Rainbow Health Victoria. The survey was run from 10 April 2021 to 3 August 2021. Given that most respondents identified as GBMSM or TGD, the decision was made to limit the analysis to only include these respondents.

### Survey Instrument

The survey was accessed through an online link (hosted by Qualtrics). The survey collected data on sociodemographic characteristics and sexual practices. Respondents were provided with a list of app/website functions and asked to rate how useful each function would be using a five-point Likert scale. Participants were also asked about current and preferred HIV/STI testing reminders and were able to rank their top three answers. Participants were not required to answer all questions and could rank less than three options if desired. Free text response to the questions: “*If you could design an app, website, or health service for LGBTIQA+ people that would make it easier to get tested for HIV/STIs? What would it do? Feel free to be creative--all answers and ideas are welcome!*” this allowed participants to expand on any features they would want in an eHealth intervention.

### Statistical Analysis

We used descriptive statistics to summarise the characteristics of the study population, using Stata (version 17, StataCorp, College Station, TX). Differences between GBMSM and TGD were assessed using Chi-squared test. The free-text responses were thematically analysed using NVivo (Release 1.6, QSR International Pty Ltd., Melbourne, Australia). Ethics approval was granted by the Alfred Ethics Committee (670/20).

## Results

The survey was accessed 727 times during the study period, and 704 people consented to participate of whom 513 (72.9%) completed the survey. There were 47 (9.2%) participants who did not identify as GBMSM or TGD and were excluded, leaving a total of 466 participants for the analysis. Most were recruited through an SMS or email from MSHC (306/466, 65.7%), followed by Grindr (93/466, 20.0%), then community groups (41/466, 8.8%).

Table 1 summarises the sociodemographic characteristics of the study population. Of the 466 participants, the majority identified as cisgender males (92.7%). The median age was 48 (interquartile range [IQR]: 37-56) and three-quarters were born in Australia. There were 21.2% (*n*=98) of participants living with HIV and had an undetectable viral load. Among those not living with HIV, 37.8% (*n*=139) reported PrEP use.

**Table 1:** Sociodemographic characteristics of the study population (N=466)

| Demographic | n (%) |
| --- | --- |
| **Current gender identity** |  |
| Cisgender Male | 432 (92.7) |
| Transgender Male | 8 (1.7) |
| Transgender Female | 1 (0.2) |
| Non-binary or gender-fluid | 21 (4.5) |
| Different identity <sup>A</sup> | 4 (0.9) |
| **Sexual identity** |  |
| Gay | 410 (88.0) |
| Bisexual | 36 (7.7) |
| Queer | 14 (3.0) |
| Lesbian | 1 (0.2) |
| Straight/Heterosexual <sup>B</sup> | 1 (0.2) |
| Other, please specify | 3 (0.6) |
| Prefer not to answer | 1 (0.2) |
| **Age group (years)** |  |
| ≤ 25 | 27 (5.8) |
| 26-35 | 70 (15.0) |
| 36-45 | 116 (24.9) |
| ≥ 46 | 253 (54.3) |
| **Country of birth** |  |
| Australia | 346 (74.3) |
| Outside of Australia | 120 (25.8) |
| **HIV status** |  |
| HIV negative | 351 (75.3) |
| Positive & undetectable | 98 (21.0) |
| I don't know my status | 13 (2.8) |
| Prefer not to answer | 4 (0.9) |
| **Reports PrEP use<sup>D</sup>** |  |
| Yes | 139 (37.8) |
| No | 225 (61.1) |
| I don't know | 2 (0.5) |
| Prefer not to answer | 2 (0.5) |

<sup>A</sup> Different gender identity included (verbatim), “Genderqueer”, “Mostly male, except on Saturday nights”, “Trans femme”, and “Trans (agender)”. <sup>B</sup> Identified as Trans or gender diverse; <sup>C</sup> Different sexual identities included: 1. Pansexual, 2. Bisexual and queer, 3. Trixix (Non-binary person who loves women) <sup>D</sup> Denominator excludes those who are HIV positive and undetectable (n=98) and is thus N=368.

### Website vs App

Overall, there was no clear preference for an app or website-based sexual health platform. Of the 462 participants that answered the question, 160 (34.6%) would use either a website or an app, 165 (32.7%) would prefer a website, 119 (25.8%) would prefer an app, and 33 (7.1%) would not use either platform. There was no significant difference between GBMSM and TGD people (Supplementary Table 1). Figures 1 and 2 provide the preferences for functions on an app and web-based platform, respectively. Supplementary Figures 1-4 separates out preferences of GBMSM and TGD, and found that GBMSM were more likely to prioritise information about pre-exposure prophylaxis (PrEP) and post-exposure prophylaxis (PEP) (Supplementary Figures 1-4).

**Figure 1:**
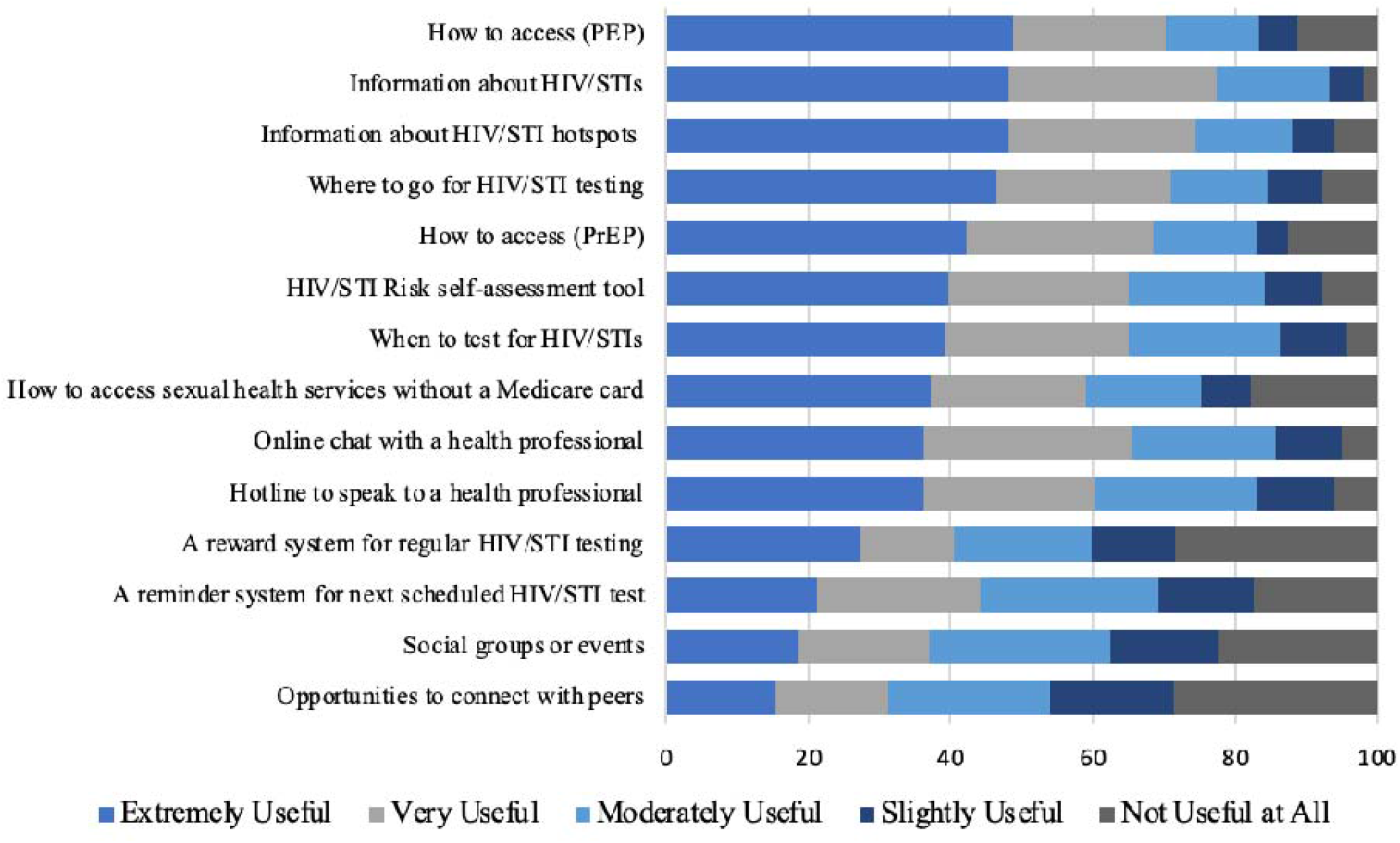
Preferences forl functions on an app-based platform among GBMSM and TGD

**Figure 2:**
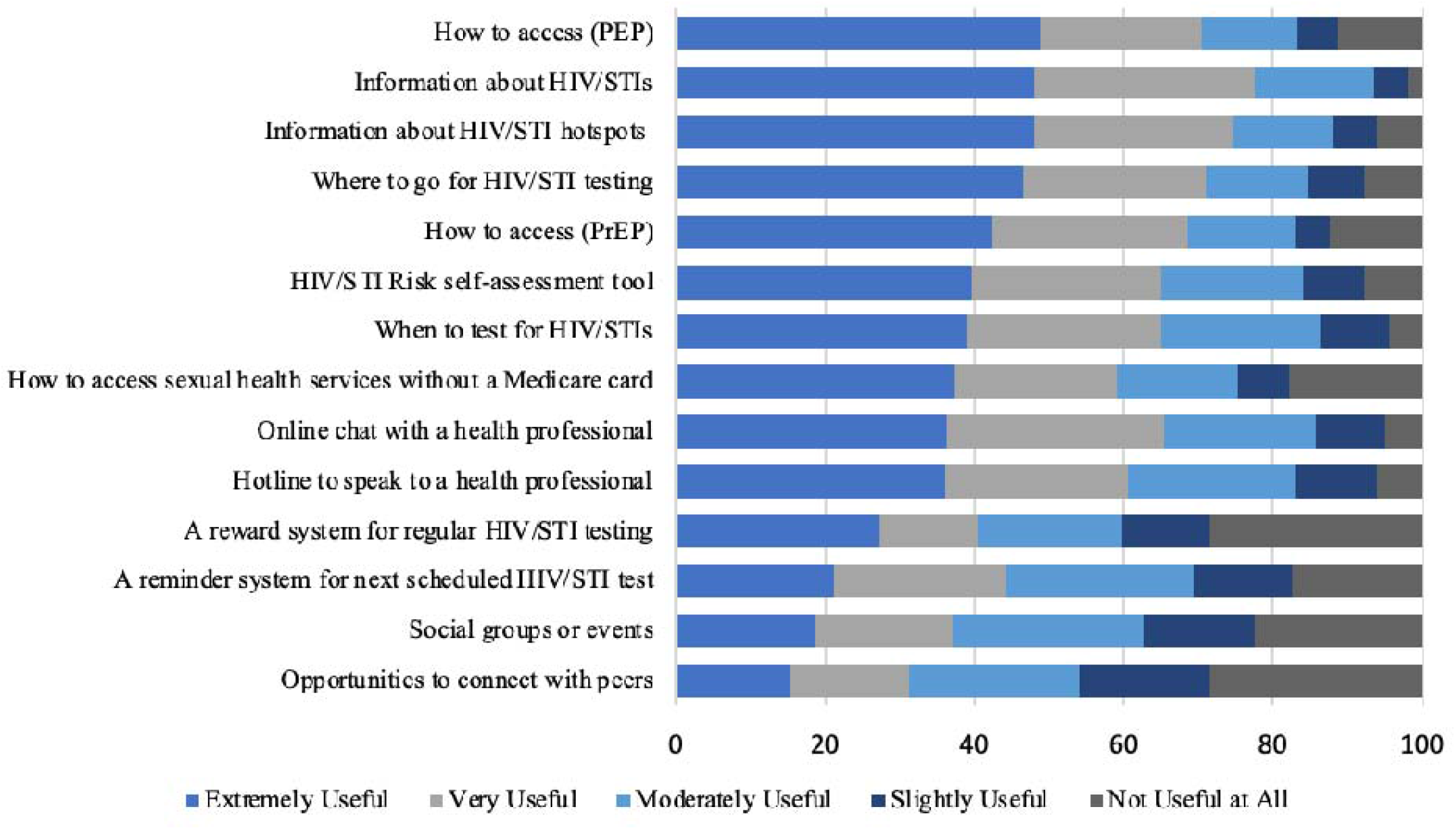
Preferences for functions on a web-based platform among GBMSM and TGD

In the free-text response, there was a strong emphasis on the need for information, particularly about STI clinics, HIV/STI hotspots (geographical areas with high levels of STI diagnoses), and sexual health education. Concerning STI clinics, respondents wanted information on their location, opening hours, contact details, and the cost of the services. Other themes that emerged included the need for anonymity, accessibility, and a simple user interface. A few participants also highlighted that they would want a discreet app. Participants were interested in features such as a sexual activity tracker (i.e., an online diary that recorded sexual encounters), a daily PrEP reminder system, and an HIV/STI testing reminder system. Some participants suggested introducing an online booking system for clinics, which would allow them to book an appointment in advance.

### HIV/STI Testing Reminder

A large proportion of respondents would prefer regular SMS reminders (112/293, 38.2%), followed by regular email reminders (55/293 18.8%) and a reminder function on their phone (48/293, 16.4%). (Figure 3) Testing reminder preferences were similar between GBMSM and TGD people (Supplementary Figures 5-6).

**Figure 3:**
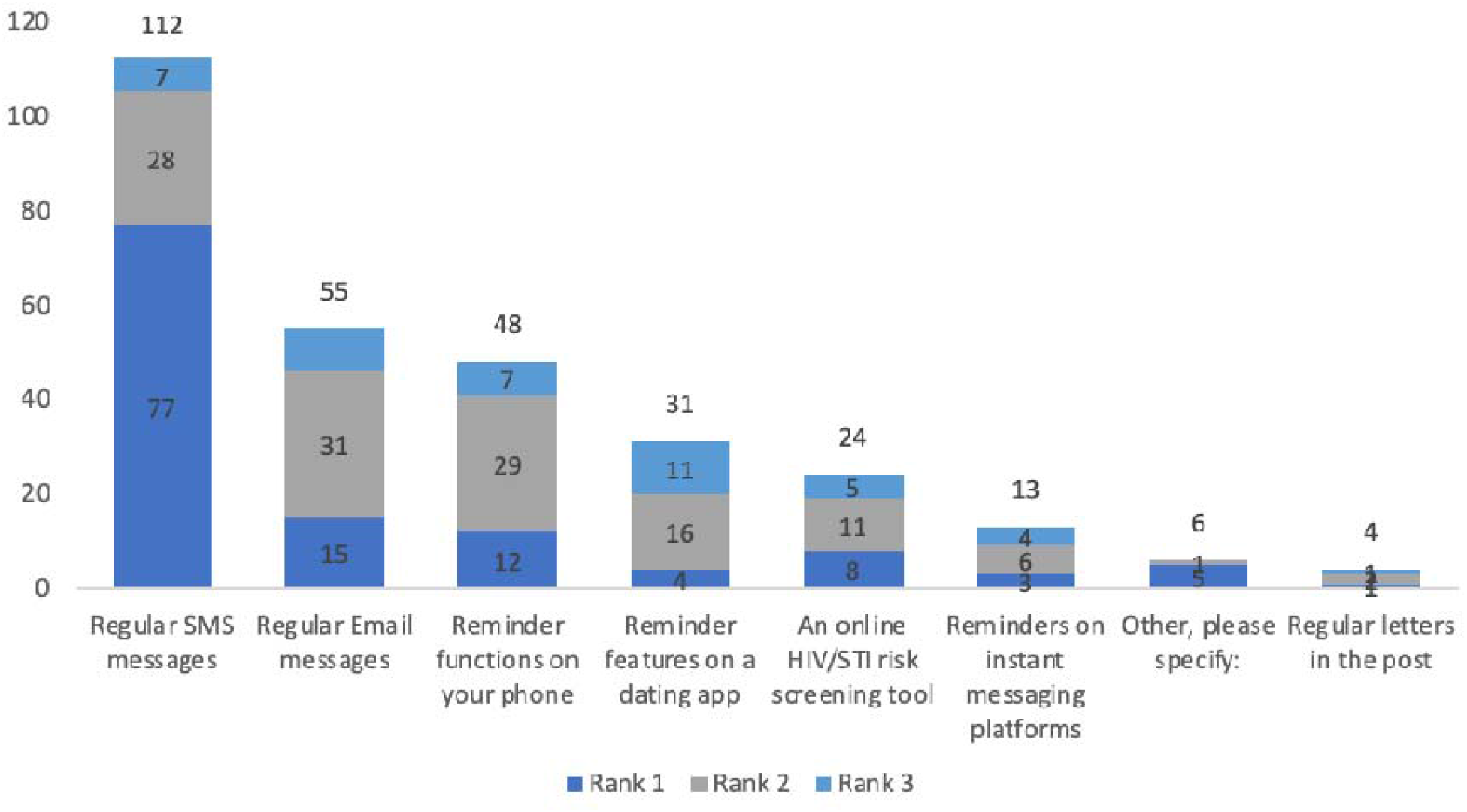
Preferred HIV/STI testing reminder system on an eHealth platform, stratified by rank

## Discussion

This study provided insights into how GBMSM and TGD people viewed the use of eHealth for sexual health. Specifically, we identified what features may be desirable for a new eHealth intervention, providing useful information for future implementation research to optimize the use of eHealth among GBMSM and TGD. Participants highlighted their desire for privacy and sexual health information. Furthermore, we found that GBMSM were more likely to prioritise wanting information about PrEP and PEP in comparison to TGD people.

The use of eHealth is a relatively new concept, and international data suggests that this has been accepted elsewhere. A study among 495 GBMSM from the USA found that 91% would be interested in an app with sexual health features tailored to GBMSM.^11^ A 2015 study using qualitative interviews of 35 MSM in China found that they supported the use of mobile phones and websites for sexual health.^12^ Most participants agreed that this provided a level of convenience, and provided information about testing services. Similarly, Nguyen et al. conducted five focus group discussions among MSM in Vietnam. The majority of participants were agreeable to eHealth. They also requested similar content to our participants, such as information about HIV/STIs and testing, and safe sex practice.^13^ The dissemination of sexual health information through social media and dating apps was acceptable to GBMSM in England, however, some participants felt that receiving this information on dating apps could negatively impact user experience.^14^ These participants felt that it may dampen the mood when searching for sexual partners, and create negative connotations with new partners.

Ensuring privacy when accessing the services was a prominent theme. It is unclear whether these concerns were due to the stigma surrounding sexual health or whether they resulted from a previous negative experience. Muessig et al. also reported concerns about privacy and confidentiality when accessing eHealth for sexual health.^12^ Another study found that the use of eHealth would depend on the privacy policy for most participants in the study.^15^ As a result, they may feel that they need to attend a specialist clinic to meet their healthcare needs. However, this can be difficult to access, and these barriers can discourage GBMSM and TGD people from seeking traditional sexual health services such as face-to-face consultations with health care professionals. eHealth addresses these barriers by offering privacy and easy access to sexual health services. These eHealth interventions can be delivered through computers, websites, and personal devices and can deliver content tailored to an individual’s needs. In turn, these address barriers to health care access like cost and accessibility.^16^ This was a major concern for respondents, and our findings suggest that future eHealth interventions should prioritise anonymity to ensure uptake and continued use of the services.

SMS for HIV/STI testing reminders was the most popular choice among respondents in our survey. Several studies have already evaluated the use of an SMS reminder system on HIV/STI testing. A minority of participants from our study indicated that they would prefer an email reminder, however, this has not been evaluated previously. A 2013 study conducted by Zou et al. at MSHC found that GBMSM who received quarterly STI testing reminders by text or email were more likely to return for a test (median 3 vs 1 test in 12 months) than controls.^17^ However, this study did not stratify results by the modality used. A 2011 study looking at STI testing rates among Australian GBMSM found that those who received 3 to 6-monthly SMS reminders were 4.4 times more likely to retest for HIV/STIs than those who did not receive a text.^18^ Similarly, a 2019 USA study reported that men who received a quarterly SMS testing reminder had a shorter interval between HIV tests than men who did not receive a text.^19^ SMS reminders are a feasible intervention that is relatively affordable and can help increase STI testing rates and has a lower burden on the health system than phone calls or other in-person interventions. Second, mobile technology is almost universally used and can effectively reach a wider population. A 2020 meta-analysis found that there were high levels of feasibility for mHealth tailored to GBMSM. However, most studies reviewed were pilot trials, and it is unclear whether these mHealth interventions would be successful on a larger scale.^20^

Our study has several limitations. First, this is a cross-sectional study, and thus we cannot make causal inferences. Secondly, our study only recruited through one gay dating app (Grindr) but no other mainstream apps, such as Tinder, Bumble, and Hinge. This may have skewed the results, and as a result, we only included GBMSM and TGD people in the analysis; thus, our findings are not generalisable to other members of the LGBTIQA+ population. To make any future intervention successful, it will require further collaboration between developers and a more representative sample of the population. Third, our sample was mostly derived from men who had recently attended a sexual health service. These individuals would therefore be biased towards individuals who are comfortable attending services and so our estimates are likely to underestimate preferences for eHealth services. Future studies should seek the views of individuals who are at risk but not attending services although we appreciate undertaking such a study is difficult. Finally, HIV/STI-related stigma, potentially influencing perceived engagement with digital platforms and data security, was not measured.

## Conclusion

Overall, our study suggests a promising future for eHealth among GBMSM and TGD people. Sexual health is still a stigmatised area, and eHealth may circumvent barriers this population faces. Further research that provides in-depth data on the themes raised in this study is required to ensure the acceptability and feasibility of any future interventions. Specifically, confidentiality and options to remain anonymous should be considered in future developments of eHealth for sexual health.

## Data Availability

All data produced in the present work are contained in the manuscript

## Acknowledgements

We thank the survey participants for contributing to the research. JO conceived the idea. EC, DL, FK, LM, JG, BB, BS, SJ, JW assisted with recruitment. EA and TP analysed the data. EA wrote the first draft of the paper. All authors contributed to writing the manuscript and approved the final version for submission. EPFC and JJO are each supported by an Australian National Health and Medical Research Council Emerging Leadership Investigator Grant (GNT1172873 and GNT1193955, respectively). CKF is supported by an Australian NHMRC Leadership Investigator Grant (GNT1172900). All other authors have no conflicts of interest to declare.

## Supplemental Data

**Supplemental Table 1.**
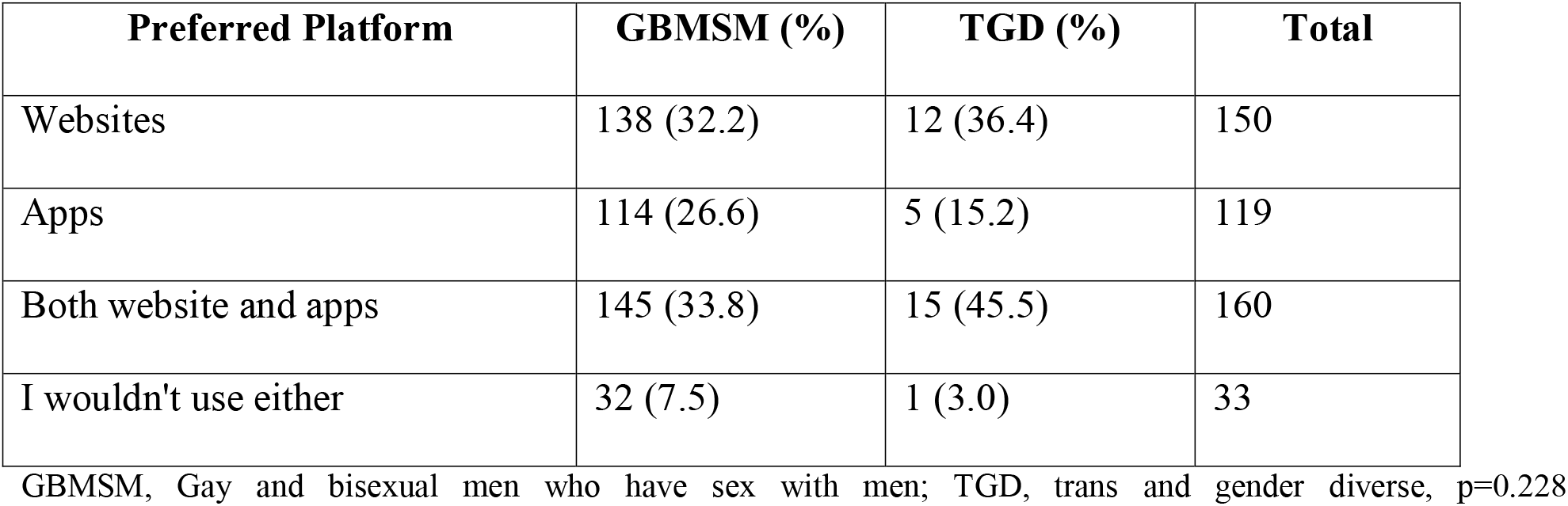
Preference for eHealth intervention platform, by risk group

| **Preferred Platform** | **GBMSM (%)** | **TGD (%)** | **Total** |
| --- | --- | --- | --- |
| Websites | 138 (32.2) | 12 (36.4) | 150 |
| Apps | 114 (26.6) | 5 (15.2) | 119 |
| Both website and apps | 145 (33.8) | 15 (45.5) | 160 |
| I wouldn't use either | 32 (7.5) | 1 (3.0) | 33 |

329 GBMSM, Gay and bisexual men who have sex with men; TGD, trans and gender diverse, p=0.228

**Supplementary Figure 1:**
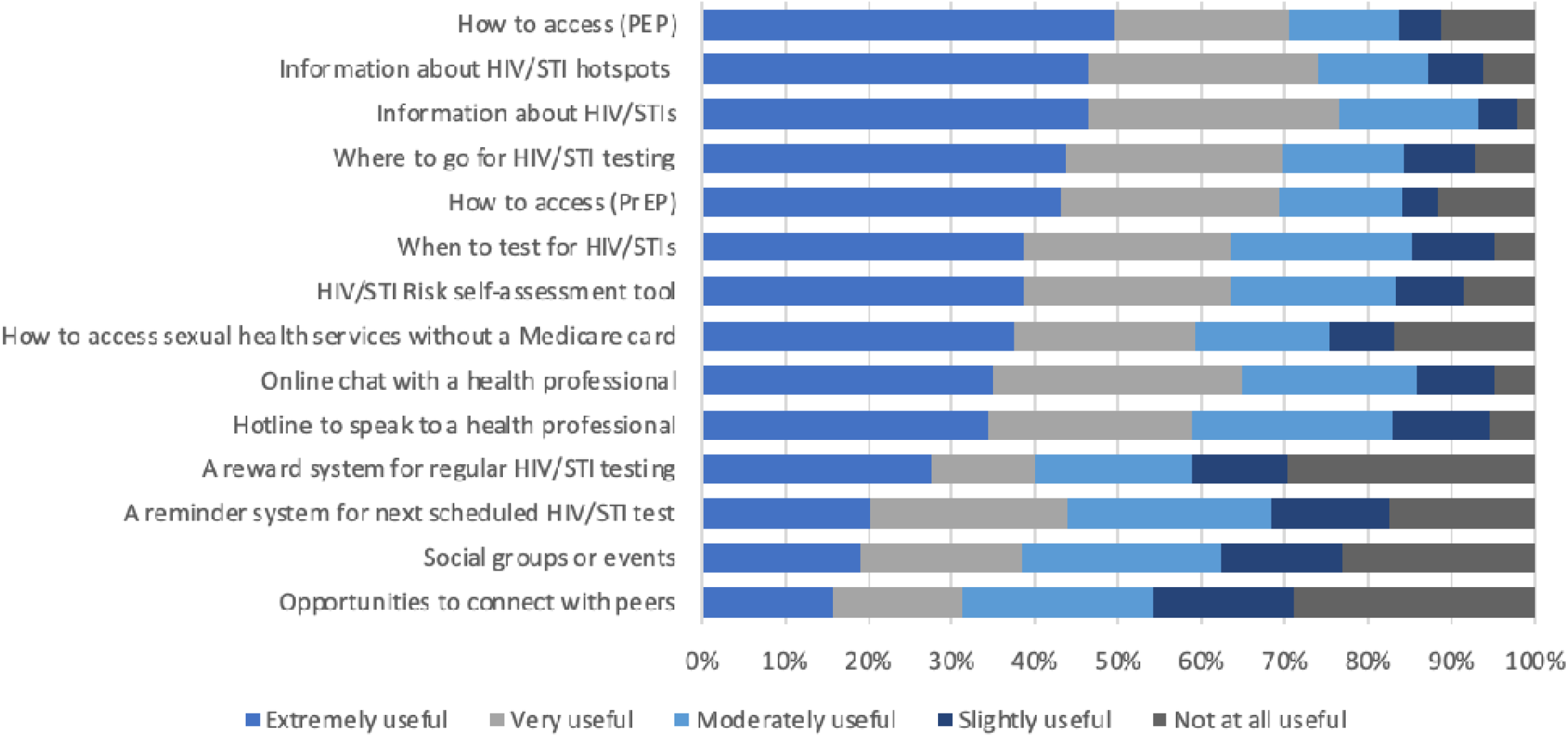
Preferences for functions on a web-based platform among GBMSM

**Supplementary Figure 2:**
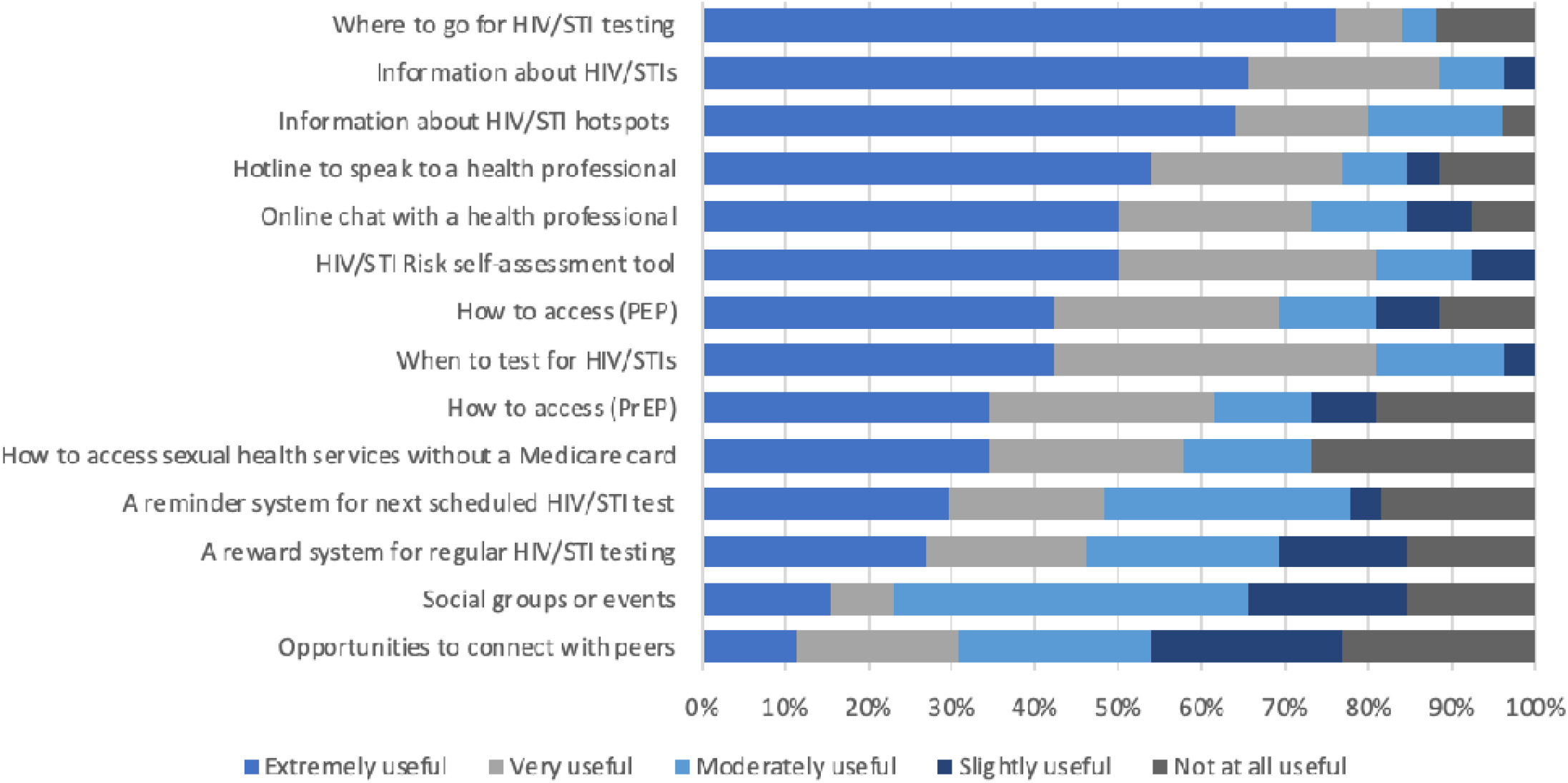
Preferences for functions on a web-based platform among TGD people

**Supplementary Figure 3:**
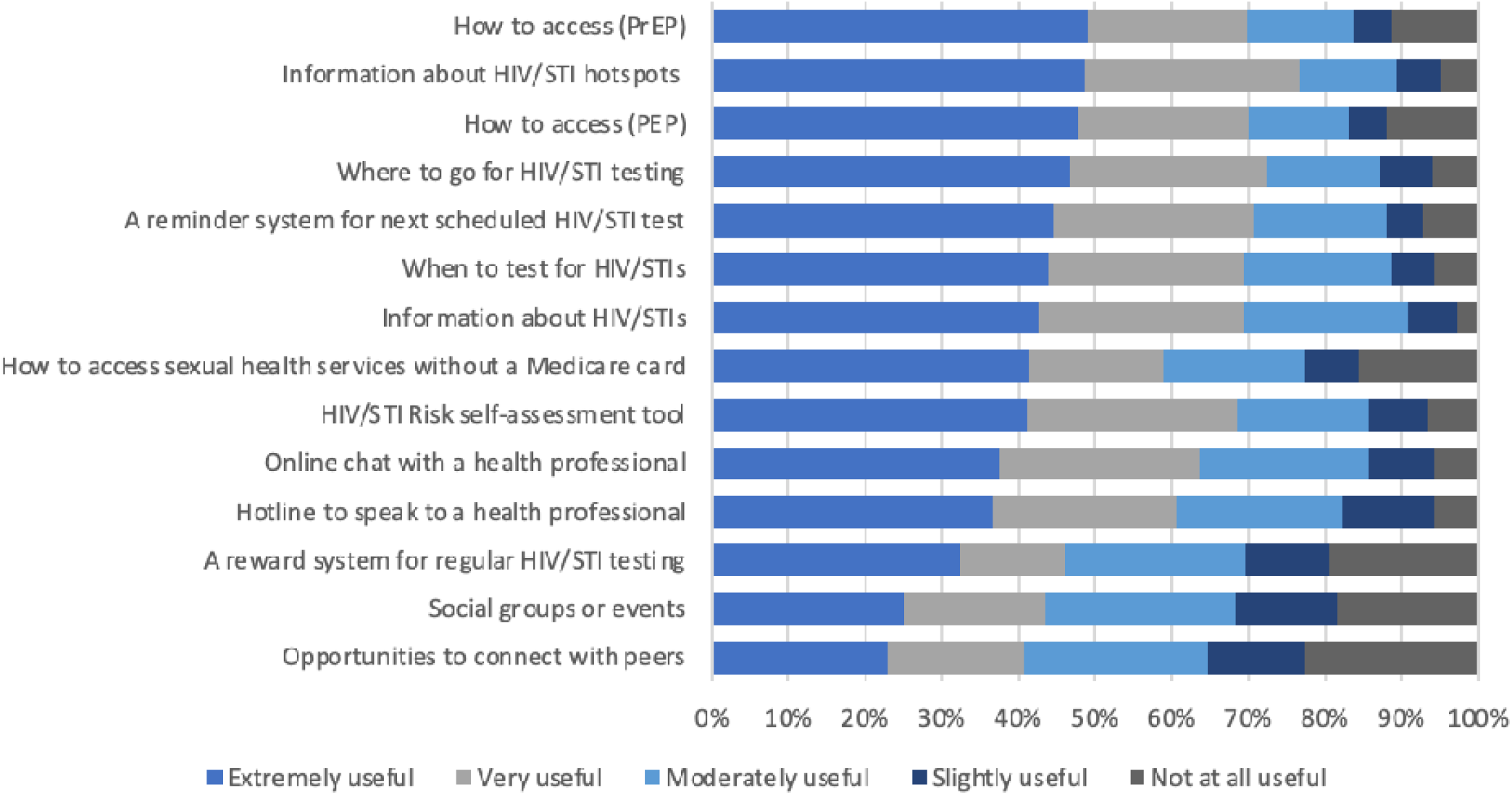
Preferences for functions on an app-based platform among GBMSM

**Supplementary Figure 4:**
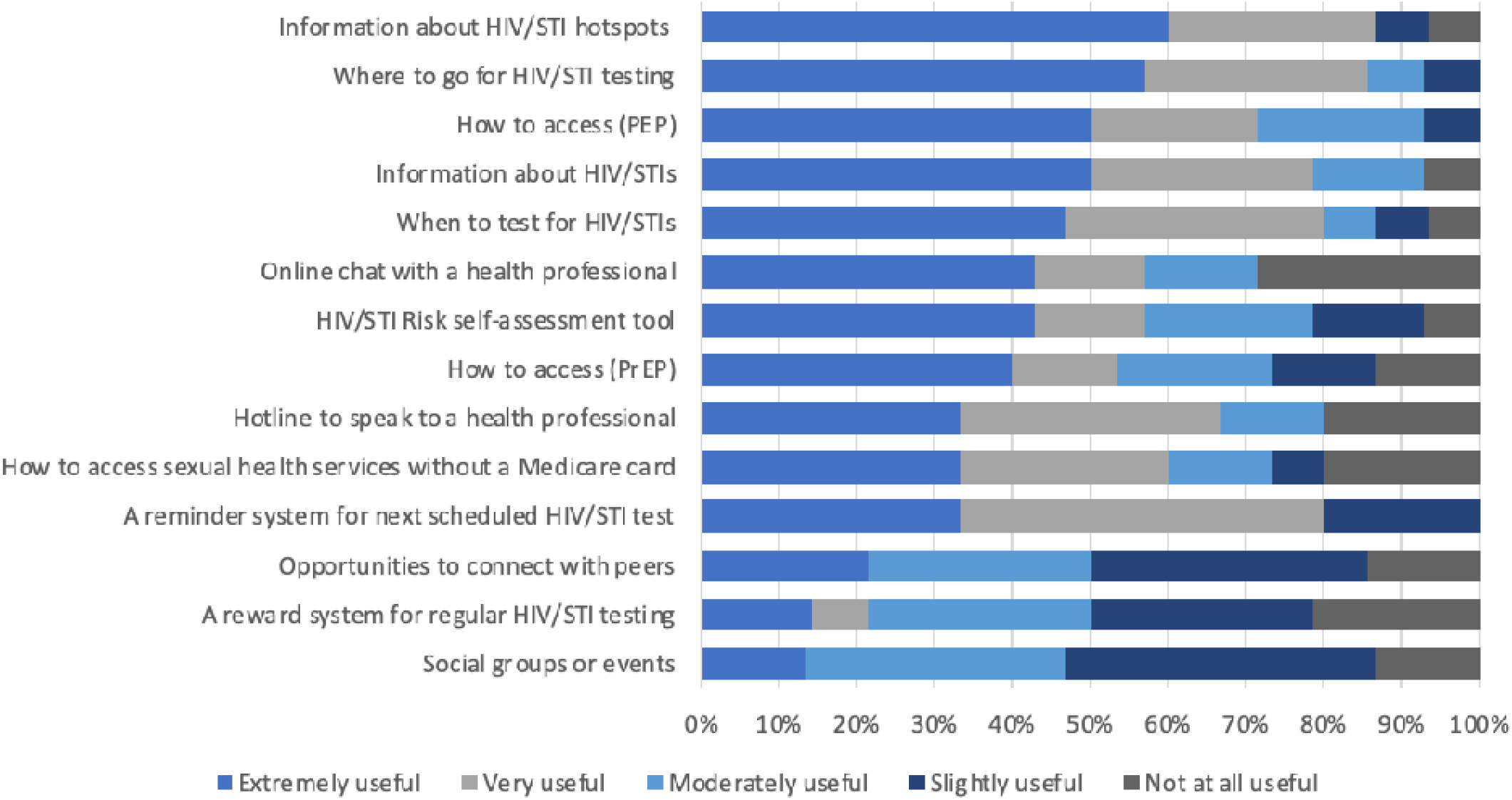
Preferences for functions on an app-based platform among TGD people

**Supplementary Figure 5:**
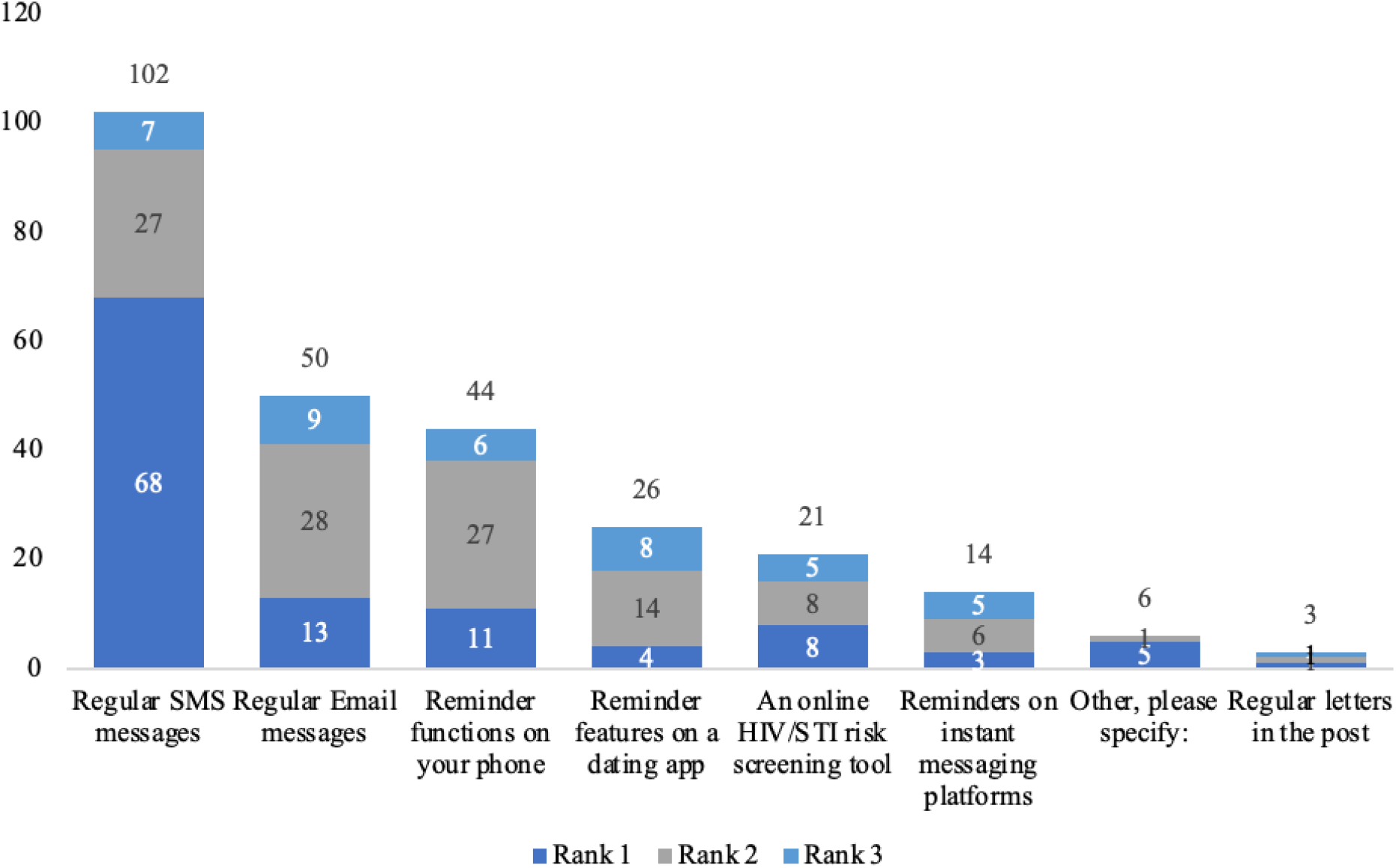
Preferred HIV/STI testing reminder system on an eHealth platform among GBMSM

**Supplementary Figure 6:**
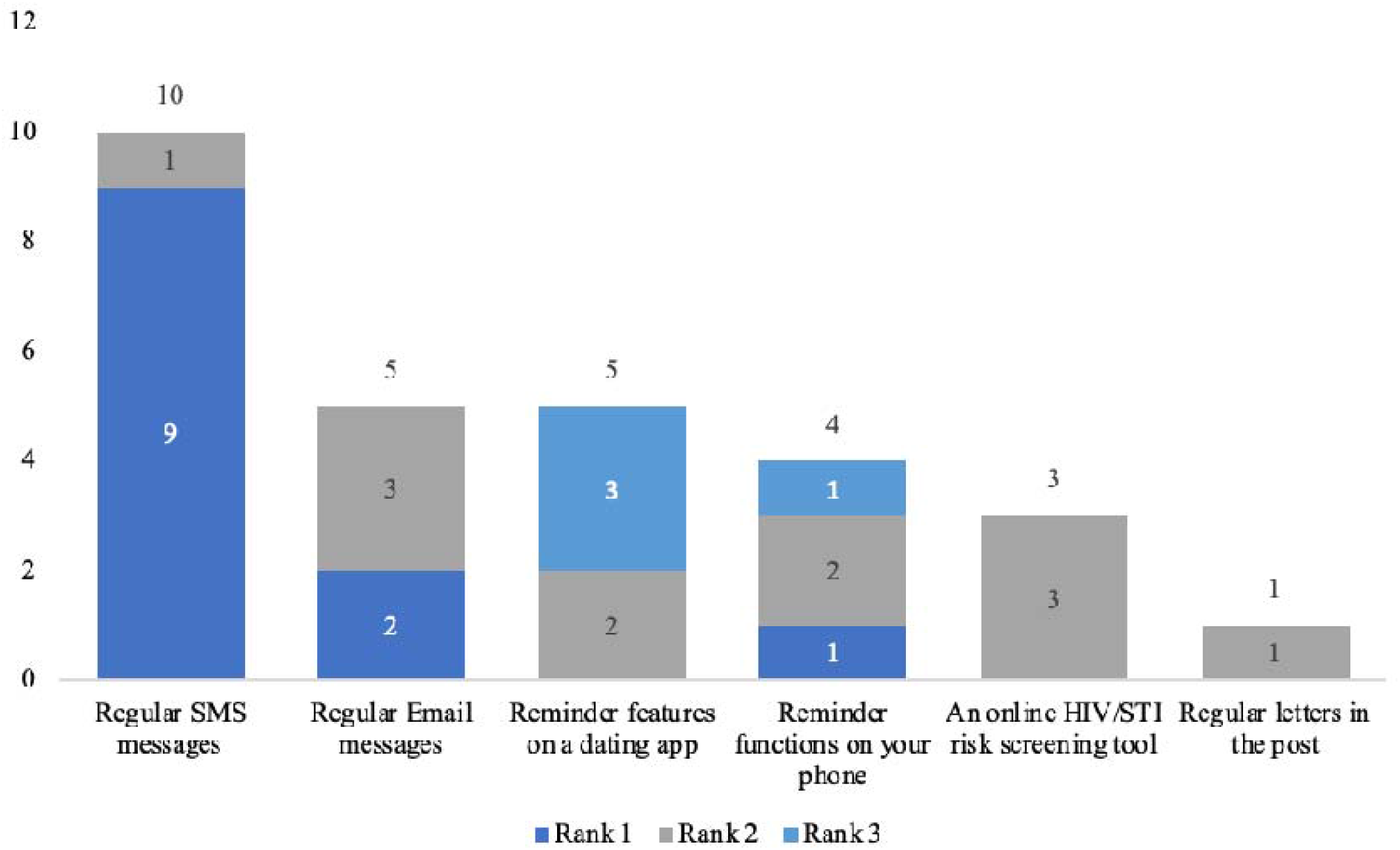
Preferred HIV/STI testing reminder system on an eHealth platform, among TGD people

